# Anxiety and sleep quality in the undergraduate stress–depression link: A moderated mediation study

**DOI:** 10.1101/2025.08.01.25332606

**Authors:** Celsa Koebeli, Marcel Koebeli, Kevin Morgan, Trudi Edginton, Laura Boubert

## Abstract

First-year undergraduate students are particularly vulnerable to stress-related anxiety and depression, and they generally exhibit poorer sleep quality compared with other student populations. While anxiety is known to explain how stress contributes to depression, its specific mediating role in the context of undergraduate-related stress remains unclear. Moreover, although poor sleep is highly prevalent in this group, its potential to exacerbate or buffer these mental health risks has been largely overlooked. To investigate these issues, this study examined whether anxiety acts as an intermediary in the link between undergraduate stress and depressive symptoms and whether sleep quality influences the intensity of these connections. This observational study included a final sample of 102 first-year psychology students (M_age_ = 19.75 ± 1.76 years) who completed assessments measuring sleep quality, stress, anxiety, and depression. Descriptive statistics, correlational analyses, and path analyses were conducted to determine the prevalence of poor sleep, variable associations, and the mediating and moderating roles of anxiety and sleep quality. Poor sleep quality was highly prevalent (61.8%) and significantly correlated with undergraduate stress (ρ = 0.42, p < .01), anxiety (ρ = 0.46, p < .01), and depression (ρ = 0.44, p < .01). Anxiety significantly mediated the relationship between undergraduate stress and depression (indirect effect = 0.23, 95% CI [0.09, 0.38]). Additionally, sleep quality moderated the anxiety–depression pathway (β = 0.04, p < .05) but not the stress–anxiety or stress–depression pathways. Anxiety plays a central role in linking undergraduate-specific stress and depression in first-year students. Crucially, poor sleep quality selectively intensified the association between anxiety and depressive symptoms, placing students with both elevated anxiety and poor sleep at high risk. Early, dual-targeted interventions may help prevent long-term depressive consequences. Future research should clarify temporal pathways and identify which stress and sleep components best predict depressive symptoms.

## Introduction

The transition from secondary education to university requires first-year students to adjust to a new environment while navigating increased independence from their families. This period of adjustment can lead to an elevated likelihood of developing mental health issues relative to peers of the same age [1, 2]. This increased vulnerability stems from the distinct pressures of university life, involving social, academic, financial, and lifestyle demands [3–7]. Collectively referred to as undergraduate stress, such pressures can increase anxiety, depression, and sleep disturbances in first-year students. The extent of this issue has been reported by the American College Health Association Survey [7], indicating that 40% of students experience high stress levels, 24% exhibit symptoms of depression, 34% experience anxiety, and 23% experience sleep difficulties. This rising concern extends across diverse cultural and economic contexts, suggesting a global public health issue [8–10].

The stress-vulnerability model by Zubin and Spring [11] provides a valuable theoretical framework for understanding university students’ mental health dynamics. This model posits that mental health issues arise from the interaction between an individual’s inherent vulnerabilities and the stressors they experience. It emphasises the need to examine the direct effects of stress and the factors that mediate and moderate its impact on psychological well-being. Examining these mechanisms is essential for designing effective interventions that mitigate mental health risks among university students.

Recent research has reinforced this framework by proposing a mediation model in which stress is associated with depression both directly and indirectly in the general population [12–15], as well as specifically among undergraduate students [16, 17]. These findings indicate that while some students develop depressive symptoms as a direct response to stress, for others, stress initially manifests as anxiety, which eventually gives rise to symptoms of depression.

Although the interplay between stress, anxiety, and depression is well-documented, sleep quality—another crucial factor influencing student well-being—has gained increased attention as a potential vulnerability. Kline defined sleep quality as a multifaceted construct encompassing subjective satisfaction with various parameters, including sleep duration, the time taken to fall asleep, and the feeling of being refreshed upon waking [18]. Notably, suboptimal sleep is a common issue among university students and appears to be particularly pronounced among first-year students [19]. Studies indicate that up to 75% of students in their first year experience suboptimal sleep [20, 21]. This prevalence is especially problematic, as poor sleep quality can impair emotion regulation [22, 23], potentially hindering students’ ability to navigate the emotionally challenging university environment during their first year [24, 25]. Consequently, poor sleep quality may threaten students’ psychological well-being and compromise their academic performance.

In light of this, the high prevalence of poor sleep among undergraduate students has prompted further investigation into its relationship with stress, anxiety, and depression, uncovering significant associations [22, 26–28]. Longitudinal studies on the sequence of these relationships suggest that stress primarily contributes to disturbed sleep [29–31], and that disturbed sleep tends to appear before signs of anxiety and depression manifest [32–34]. Considering these established associations, multiple cross-sectional studies have investigated their relevance in higher education. For instance, research on university students found that sleep disturbances helped explain how stress relates to both anxiety [35] and depression [36], suggesting that sleep problems may be a key pathway linking stress to mental health difficulties in this population. More recently, Borges, Ellis, and Ruivo Marques [37] revealed that sleep effort—the conscious attempt to achieve restful sleep, often associated with insomnia [38]— may underlie the connection between anxiety and depression in students enrolled in higher education. Consequently, students who overexert themselves to fall asleep may paradoxically remain awake longer, which can exacerbate emotional distress and help explain the well-established relationship between anxiety and depression.

Additionally, poor sleep helps explain how stress contributes to anxiety and depression and may clarify the well-established relationship between the two conditions. However, these studies adopted cross-sectional designs, which precludes causal inference. Additionally, sleep quality is most often examined solely as a mediator—that is, as a variable that explains the mechanism of an effect—leaving individual differences in vulnerability to stress-related mental health problems unaccounted for.

A methodological alternative for examining differential vulnerability is the moderation model. Rather than focusing on explanatory mechanisms, moderation considers how the strength of associations between variables may vary depending on a third factor [39]. A key advantage of moderation analysis is that it does not require temporal sequencing, making it a valuable approach for identifying potential risk factors in cross-sectional studies. In this context, examining sleep quality as a moderator—defined as a variable that influences the strength of the relationships between stress, anxiety, and depression—may help to more precisely identify individuals who are particularly at risk.

Despite the theoretical relevance of this approach, it has rarely been examined empirically. Only a few studies have expanded on sequential models by identifying sleep quality as a key factor that amplifies the impact of stress and anxiety on the development of depressive symptoms. Specifically, Leggett et al.’s longitudinal research demonstrated that in a general adult population, disturbed sleep significantly amplified the impact of stress on depressive symptoms [40]. Similarly, Park et al. conducted two distinct moderation analyses to assess sleep quality as a moderator of both stress-related anxiety and depression [41]. Consistent with the findings of Leggett et al. stressed individuals with poor sleep quality face an increased risk of developing depression [40]. However, no such effect was found for the stress–anxiety relationship. These findings are important, as they suggest that poor sleep quality may selectively intensify certain psychological outcomes of stress—namely, depressive symptoms—while leaving others, such as anxiety, unaffected. Yet, a key limitation of both studies is that they focused solely on the direct relationship between stress and depression, without considering anxiety as a mediator—an indirect pathway that has been supported in studies unrelated to sleep quality [12–16].

This omission is critical because the inclusion of a mediator, such as anxiety, can sometimes nullify a previously significant moderation effect by accounting for variance in the outcome variable [42]. Consequently, failing to incorporate the indirect pathway through anxiety risks oversimplifying how stress, sleep quality, and depression are interrelated, particularly in moderation models that assess sleep as a risk factor. This may lead to misinterpretations and compromise the validity of such models. To address this limitation, models that integrate both unmediated and mediated pathways are required to better capture the complexity of the stress–depression relationship.

Ho employed this approach to investigate how stress, sleep quality, and psychological outcomes, such as anxiety and depression, interact within a sample of Facebook users holding administrative roles [43]. The author employed a moderated mediation model that included both a direct link from stress to depression and an indirect effect through anxiety, with sleep quality moderating the latter part of the indirect pathway. The findings revealed that Facebook users who reported reduced sleep quality and concurrent heightened anxiety levels had an elevated risk of depression. However, the interpretation of these findings requires careful consideration for several reasons. First, the sample presents limitations: this study lacked a control group of non-Facebook users, making it unclear whether the findings apply only to Facebook users or extend to a broader population. Additionally, the sample was described as belonging to an administrative body, but no further details were provided, leaving its characteristics ambiguous. Second, Ho’s [43] model limits the role of sleep quality to moderating the link between anxiety and depression, failing to consider its potential effects on the direct association between stress and depression, as well as its impact on the early stage of the indirect stress-to-anxiety pathway [44]. Since earlier studies have shown that sleep quality can alter the strength of the link between stress and depressive symptoms, typically examined through basic interaction models [40, 41], Ho’s model [43] could have taken the opportunity to provide deeper insights by extending these findings within a more complex moderated mediation framework. Moreover, such a model would have provided an opportunity to pinpoint where sleep quality exerts its strongest influence—an insight that is essential for developing more targeted intervention strategies.

For example, if sleep quality amplifies the unmediated pathway (stress to depressive symptoms) or the first leg of the mediated pathway (stress-to-anxiety), interventions should focus on improving sleep quality as a preventive measure against both anxiety and depression. Conversely, if the strongest moderation effect occurs in the latter part of the mediated pathway (from anxiety to depressive symptoms), interventions should prioritise addressing poor sleep quality in individuals with anxiety to prevent its escalation into depression.

Consequently, examining the role of sleep quality beyond isolated pathways— considering its moderating effect on both the direct stress–depression relationship and the complete indirect pathway through anxiety—could yield enhanced insight into how it affects stress-related emotional outcomes [44]. This approach could refine previous findings [40, 41, 43], as the direct stress–depression relationship may lose significance when sleep quality moderates all paths in the model.

Building on these considerations, this study, grounded in the stress-vulnerability model [11], used a moderated mediation design to investigate how sleep quality moderates all pathways connecting undergraduate stress-to-anxiety and depressive symptoms in first-year students. By specifically focusing on undergraduate stress, it aids in modelling the unique stressors experienced by university students and their distinct effects on mental health. This approach extends previous research and provides a framework for developing more targeted prevention and treatment strategies.

Specifically, this study aims to (a) determine the extent of subjective poor sleep quality among first-year undergraduate students; (b) validate the associations among sleep quality, undergraduate stress, anxiety, and depressive symptoms; (c) ascertain whether anxiety serves as a mediator in the nexus between undergraduate stress and depression; and (d) evaluate the impact of inadequate sleep quality as a possible risk factor for stress-induced anxiety and depression among undergraduates.

Based on previous research, the following hypotheses have been formulated:

1. Poor sleep quality is highly prevalent among first-year undergraduate students.
2. Students experiencing poor sleep tend to report more undergraduate-specific life stressors, as well as symptoms of anxiety and depression.
3. Anxiety helps explain the association between undergraduate stress and depression.
4. Sleep quality moderates (a) the direct pathway from undergraduate stress to depressive symptoms and (b) the second segment of the indirect path: the link between anxiety and depression.
5. No moderating role of sleep quality is anticipated in the association between undergraduate stress and anxiety (i.e. the first segment of the indirect path).

## Materials and methods

### Study design and setting

This cross-sectional study was conducted at the University of Westminster’s Psychology Research Lab, located on New Cavendish Street, London. Data were collected between January 6^th^ 2014 and June 2^nd^ 2017 as part of the first author’s PhD research. Participants were recruited through the university’s SONA Research Participation Scheme. During a single session, they completed a series of validated self-report questionnaires assessing variables such as anxiety, depression, sleep quality, and undergraduate stress. Notably, there were no follow-up periods or specific exposures involved in this study. Approval for this research was granted by the Ethics Committee of the University of Westminster (reference number: VRE1314-1045). Although data collection occurred several years ago, the relationships among the study variables have been consistently supported in research from 2008 to 2022 [12, 13, 15, 16, 32–34, 45, 46], rendering the findings relevant and timely.

### Participants

A total of 110 first-year undergraduate psychology students at the University of Westminster participated in this study. Only students aged 18 to 25 years were included to ensure developmental consistency and align with the emerging adulthood population, as defined by Arnett [3]. Only those without a diagnosed mental health condition were included to examine subclinical psychological patterns and adjustment processes typical of the general student population, without the confounding effects of clinical diagnoses.

Eight participants were excluded owing to incomplete responses on the sleep quality measure, resulting in a final sample of 102 students (79 women and 23 men), aged 19–25 years. To minimise potential sources of bias, all questionnaires were completed anonymously in a single session. Participants were assured of the confidentiality of their responses to reduce self-report and social desirability bias. Only participants who provided complete data on all key variables were retained for the final analysis.

### Procedure

After providing written informed consent, participants completed a set of validated self-report questionnaires in a single, in-person session at the University of Westminster’s Psychology Research Lab. The questionnaires assessed anxiety, depression, sleep quality, and stress specific to undergraduates. Participation was voluntary, and data collection was conducted anonymously to reduce social desirability and self-report bias. The session took approximately 50 minutes.

### Measures

#### Sleep quality

Sleep quality was assessed by means of the Pittsburgh Sleep Quality Index (PSQI) [47], a widely used self-report questionnaire that evaluates sleep quality over the preceding month. The PSQI combines Likert-type scales with open-ended questions, allowing for a comprehensive assessment within 5–10 minutes. Through its assessment of seven key domains, the PSQI provides an overall score, with scores exceeding five indicating suboptimal sleep quality among undergraduate students [48]. The instrument demonstrates strong psychometric properties, with an internal consistency of Cronbach’s α = 0.83 and a test-retest reliability of r = 0.85 [47]. Its extensive application across diverse populations, including undergraduate cohorts [49, 50], along with its robust psychometric properties [51, 52], solidifies its position as a cornerstone instrument in sleep research.

#### Undergraduate stress

The Undergraduate Stress Questionnaire (USQ), created by Crandall et al. [53], was used to evaluate the stressors unique to undergraduate students. This 82-item checklist captures a broad spectrum of stressors. Total scores range from 0–82, with larger values indicating more intense undergraduate stress. According to Crandall et al.’s [53] original study, the USQ demonstrates satisfactory psychometric properties, including an internal consistency of KR20 = 0.8 [54], a split-half reliability of r = 0.71, and a test-retest reliability of r = 0.59 over a six-week period. Its specific focus on undergraduate stressors makes it particularly suitable for assessing stress among undergraduates in this population.

#### Anxiety

Students’ anxiety was evaluated through the State Anxiety section of the State-Trait Anxiety Inventory (STAI) [55]. Participants rated their feelings on this 20-item measure using a four-point Likert scale, generating total scores that could range from 20–80. Scores between 41 and 60 suggest moderate anxiety, while scores above 61 indicate severe anxiety [56].

The STAI demonstrates high internal consistency, with Cronbach’s α coefficients reported to be between 0.86 and 0.95 for state anxiety [55, 57]. Its construct validity and relevance to student populations have been well-established [58–60], making it an optimal choice for assessing anxiety levels in undergraduate students.

#### Depression

The Beck Depression Inventory-II (BDI-II) was used to evaluate students’ depressive symptoms [61]. This self-report instrument, comprising 21 items, assesses the extent, strength, and profundity of depressive symptoms within a two-week timeframe. This instrument produces scores between 0 and 63, with higher values reflecting more intense depressive symptomatology. The psychometric evaluation of the BDI-II demonstrates high internal consistency (Cronbach’s α ranging from 0.89–0.92) and a test-retest reliability over one week of (0.93) [61–63]. Its widespread adoption in undergraduate research [64–66] and strong validity in non-clinical settings [67] support its appropriateness for this study.

#### Covariates

Demographic variables, including age and sex, were recorded. Substance use— specifically average daily nicotine intake (estimated by cigarette count) and caffeine consumption (measured by the number of caffeinated drinks consumed daily) —was assessed as a potential confounding variable owing to its potential impact on sleep quality and mental health outcomes [68–72].

#### Definition of study variables

In line with STROBE reporting guidelines, undergraduate stress was the primary predictor variable, assessed using the USQ, while depressive symptoms (BDI-II scores) constituted the primary outcome variable. Anxiety symptoms (STAI-State scores) were utilised as a mediator, while sleep quality (PSQI global score) served as an effect modifier. Age, sex, and substance use (nicotine and caffeine) were included as potential confounders. As this study focused on symptom levels in a non-clinical undergraduate population, no formal diagnostic criteria were applied.

### Data analysis

Data analysis included descriptive statistics, non-parametric correlations, and regression-based conditional models. The statistical tests were performed using IBM SPSS version 30.0, supplemented with the PROCESS macro version 4.3 [73]. As noted in the participants’ section, eight participants with missing data on the sleep quality measure were excluded, yielding an analytic sample of 102 individuals. Total questionnaire scores (STAI, BDI, USQ, PSQI) were used as continuous variables in their original form without transformation or categorisation. Confounders such as age, nicotine use, and caffeine consumption were entered as continuous variables, while sex was included as a categorical variable (0 = male, 1 = female).

### Assumption testing for conditional modelling

First, the assumptions required for the use of Ordinary Least Squares–based estimation methods were evaluated: continuity of variables, absence of autocorrelation, linearity of relationships, homoscedasticity, independence of residuals, absence of strong multicollinearity, and the presence of observations with unusual influence. All causally affected measures (STAI, BDI) were continuous variables. Given the cross-sectional research design and the non-hierarchical data structure, it was assumed that the observations were not autocorrelated. The linearity of the relationships between the main predictions (USQ, STAI) and the outcome variable (BDI) was assessed with LOESS lines in bivariate scatter plots. The LOESS lines for the USQ-BDI and STAI-BDI relationships appeared to be linearly increasing. While the USQ-STAI relationship revealed an initially slightly negative slope followed by a positive one, it still appeared reasonable to classify it as linear overall. Robust standard errors were generated using the heteroscedasticity-consistent inference method (HC3) available in PROCESS (HC method) [74]. Scatter plots and histograms confirmed that the standardised residuals appeared randomly dispersed around zero, with only a few observations outside the ±2 SD range. This supports the assumptions of linear relationships and homoscedasticity across all models used. Variance inflation factors (VIFs), condition indices, and variance proportions were calculated to assess multicollinearity between predictor variables. Since PROCESS does not offer assumption testing, the moderated mediation models were rebuilt and analysed in SPSS as multiple regression models using mean-centred interaction terms as predictor variables. The mean-centring of the interaction terms was required to avoid inflated VIFs due to overlap with their component variables [73]. None of the VIFs exceeded the commonly acknowledged threshold of 10 for strong multicollinearity [75]. Only the VIFs for PSQI and the STAI-PSQI interaction term in Model 14, as well as the VIFs for PSQI and the USQ-PSQI interaction term in Model 59, exceeded 1.5. In all models, the condition indices remained below 20. Possible moderate collinearity was detected between PSQI and the STAI-PSQI interaction term (Model 14: inflation index of 16.558, variance proportions of 0.719 for PSQI and 0.763 for the interaction term; Model 59: inflation index of 19.128, with 0.785 for PSQI and 0.825 for the interaction term). Cook’s distances were calculated using the rebuilt regression models to detect observations with unusual influence on the regression parameters. None of the models showed a Cook’s D above 0.5, a commonly accepted threshold value. A post-hoc power analysis (G*Power 3.1.9.7) confirmed power values of 1.000 for all models, primarily due to R-squared values above 0.6, which resulted in effect sizes of 1.5 or higher.

All analyses in PROCESS were conducted using bootstrap resampling with 10,000 iterations, which provides additional robustness against potential non-normality [73]. Diagnostic analyses of all regression assumptions are presented in (S1 File).

### Mediation and moderated mediation analyses

A mediation model was tested using PROCESS Model 4, where undergraduate stress served as the predictor variable, anxiety functioned as the mediating variable, and depression was treated as the outcome variable. Following this, a moderated mediation analysis was conducted using PROCESS Model 59 to examine the modifying effect of sleep quality on all pathways linking undergraduate stress, anxiety, and depression.

To account for potential model complexity relative to sample size, three separate moderated mediation analyses were conducted using PROCESS Models 5, 7, and 14. These tested the moderating influence of sleep quality on (a) the direct effect of undergraduate stress on depression, (b) the pathway linking undergraduate stress-to-anxiety, and (c) the connection between anxiety and depression. In all conditional PROCESS Models—specifically Models 5, 7, 14, and 59—the PSI total score was entered as a continuous moderator to determine whether the strength of the stress–depression pathway, mediated by anxiety, varied as a function of perceived poor sleep quality. Statistical significance was determined based on 95% confidence intervals that excluded zero.

### Reporting guidelines

This study follows the STROBE (Strengthening the Reporting of Observational Studies in Epidemiology) guidelines for cross-sectional studies [76], as detailed in (S2 File).

## Results

### Sample characteristics

The final study sample comprised 102 undergraduate psychology students enrolled at the University of Westminster. Their average age was 19.7 years (SD = 1.76, range: 18–25), and 77.5% identified as female, reflecting the sample’s overall sex composition.

### Hypothesis 1

Hypothesis 1 proposed that a high proportion of students experience poor sleep quality. On average, participants rated their sleep quality at 7.08 (SD = 3.62). The data showed a right-skewed distribution (standardised skewness = 3.42) with normal kurtosis. A total of 61.8% of participants had PSQI scores exceeding the standard cutoff of five indicating reduced sleep quality. Consequently, the null hypothesis was rejected, supporting the conclusion that disturbed sleep was prevalent within this sample.

### Hypothesis 2

Hypothesis 2 posited strong associations among sleep quality, undergraduate stress, anxiety, and depression. Undergraduate stress showed a right-skewed distribution (standardised skewness = 2.46) with normal kurtosis. Anxiety conforms to a normal distribution. Depression exhibited a strong right skew (standardised skewness = 5.6) with elevated kurtosis (standardised kurtosis = 4.30). Given the skewed distribution of depression and several other variables, Spearman’s rank-order correlation coefficients (ρ) were used. As anticipated, lower sleep quality was significantly associated with more pronounced undergraduate stress (ρ = .418, p < .01), anxiety (ρ = .462, p < .01), and depression (ρ = .441, p < .01). These results support the hypothesised relationships, leading to the rejection of the null hypothesis (Table 1).

**Table 1.** Descriptive Statistics and Spearman’s Rank-order Correlations Among Study Variables.

|  | M | SD | 1 | 2 | 3 | 4 | 5 | 6 |
| --- | --- | --- | --- | --- | --- | --- | --- | --- |
| 1. Undergraduate stress | 18.81 | 10.41 |  |  |  |  |  |  |
| 2. Depression | 13.38 | 9.79 | 0.409** |  |  |  |  |  |
| 3. Anxiety | 41.69 | 10.51 | 0.283** | 0.682** |  |  |  |  |
| 4. Sleep quality | 7.08 | 3.62 | 0.418** | 0.441** | 0.462** |  |  |  |
| 5. Nicotine consumption | 1.46 | 3.68 | 0.077 | 0.021 | -0.004 | 0.070 |  |  |
| 6. Caffeine consumption | 2.20 | 2.51 | 0.080 | 0.024 | 0.131 | 0.028 | 0.193 |  |
| 7. Age (years) | 19.75 | 1.76 | 0.181 | 0.167 | 0.180 | 0.151 | 0.032 | 0.133 |

Spearman's $\rho$ was used. $p < .01$ (two-tailed). M = mean; SD = standard deviation.
No incomplete data were present in the final sample.

### Hypothesis 3

Hypothesis 3 proposed that anxiety acts as a partial mediator in the association between undergraduate stress and depression. PROCESS Model 4 was used to examine anxiety’s role in the association between undergraduate stress and depressive symptoms. The total effect of undergraduate stress on depressive symptomatology was 0.385, comprising a direct effect of 0.157 (95% CI [0.015, 0.299]) in the undergraduate stress–depression relationship and an indirect effect of 0.228 (95% CI [0.090, 0.377]) in the undergraduate stress–anxiety–depression relationship. Each pathway in this model, encompassing direct effects, indirect effects, and all segments of the mediated path, showed statistical significance. R-squared value for the model was 0.599, indicating that undergraduate stress and anxiety together explained 59% of the variance. Consequently, the null hypothesis (H₀) was rejected, supporting the role of anxiety as a mediator.

Neither age nor sex qualified as covariates in this analysis. Age showed no significant correlation with depression. Regarding sex differences, a Mann–Whitney U test indicated that women (N = 79, sum of ranks = 4,246) exhibited a higher mean rank compared to men (N = 23, sum of ranks = 1,007); however, this variation was not significant at the statistical level (p = .05). Nicotine and caffeine consumption were also examined as potential covariates. However, neither variable showed significant correlations with depressive symptoms (nicotine: r = 0.021, p = .832; caffeine: r = 0.024, p = .809), nor did they significantly alter the associations between the main variables when included in the models. Consequently, to maintain model parsimony, these variables were not included in further analyses.

### Hypotheses 4 and 5

Hypothesis 4 proposed that sleep quality moderates a) the associations between anxiety and depressive symptoms and b) the direct relationship between undergraduate stress and depression. Hypothesis 5 expected no moderating effect on the anxiety–depression pathway, based on previous findings [41]. Given the complexity of the model and the limited dataset, two approaches were used to test these hypotheses: 1) an initial analysis using a single moderated mediation model to examine all three moderation effects simultaneously, and 2) a secondary analysis using separate moderated mediation models to assess each moderation effect individually.

### Initial moderated mediation analysis

According to PROCESS Model 59, sleep quality did not have a significant moderating influence on any of the pathways in the proposed moderated mediation model. No significant effects emerged at any of the examined sleep quality levels (−1 SD, mean, +1 SD). The absence of significant effects may be attributable to the modest sample size (N = 102), which might not have been large enough to support the model’s complexity.

The effect sizes for the conditional direct effects of undergraduate stress on depression were 0.128, 95% CI [-0.001, 0.274] (with sleep quality at the mean level); 0.117, 95% CI [-0.018, 0.274] (with sleep quality at –1 SD); and 0.133, 95% CI [-0.120, 0.317] (with sleep quality at +1 SD). The R-squared for this model was 0.600. A significant unmoderated direct effect was observed, β = 0.157, 95% CI [0.015, 0.299]. As none of the interaction terms were significant, the null hypothesis remained supported, suggesting that sleep quality did not moderate any of the pathways in the proposed mediation model.

### Secondary moderated mediation analysis

Given the likelihood that the complexity of the full model—incorporating three interaction terms—may have limited statistical power, follow-up analyses were conducted with PROCESS Models 5, 7, and 14 to assess each moderation effect on each path independently. In Model 5, sleep quality did not significantly moderate the direct path from undergraduate stress to depression. The lack of a significant interaction led to the retention of the null hypothesis. Similarly, sleep quality did not significantly moderate the path from undergraduate stress-to-anxiety (Model 7), as this effect was also not significant, which aligns with the hypothesis of no moderation. Unlike Models 5 and 7, Model 14 revealed a strong and significant moderating role of sleep quality in the link between anxiety and depressive symptoms across all levels examined. An interaction effect of 0.037 was observed. At average sleep quality, the conditional indirect effect was 0.213, 95% CI [0.081, 0.363]; at −1 SD, it was 0.167, 95% CI [0.059, 0.302]; and at +1 SD, it reached 0.260, 95% CI [0.098, 0.440]. When moderation was not accounted for, the direct effect remained significant at 0.140, 95% CI [0.004, 0.276] (Figs 1 and 2). The R-squared for this model was 0.629. These findings confirmed the moderating role of sleep quality on the second segment of the indirect pathway and consequently led to the rejection of the null hypothesis.

**Fig 1.**
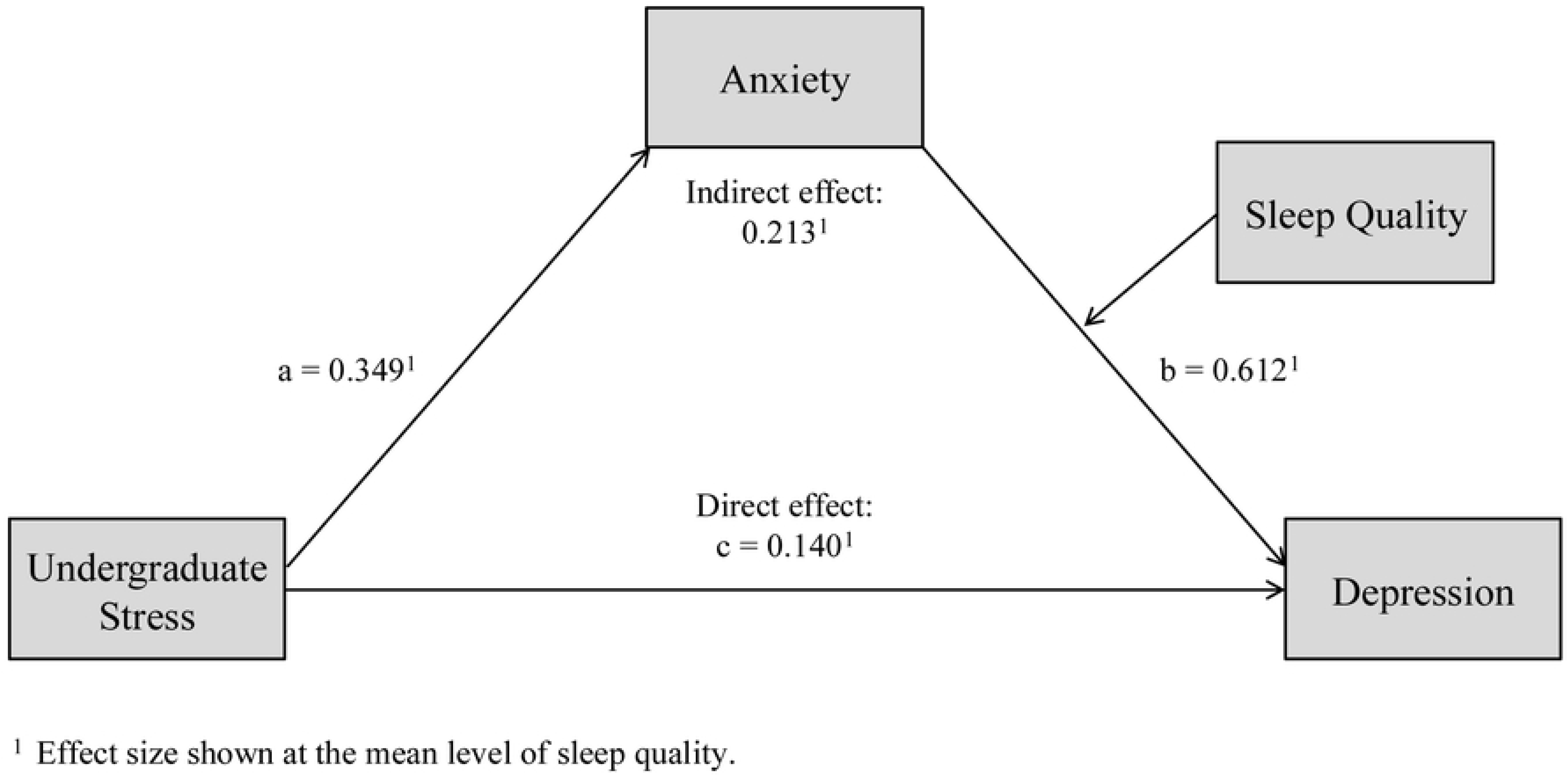
Moderated Mediation Analysis Examining the Stress–Anxiety–Depression Relationship (PROCESS Model 14)

**Fig 2.**
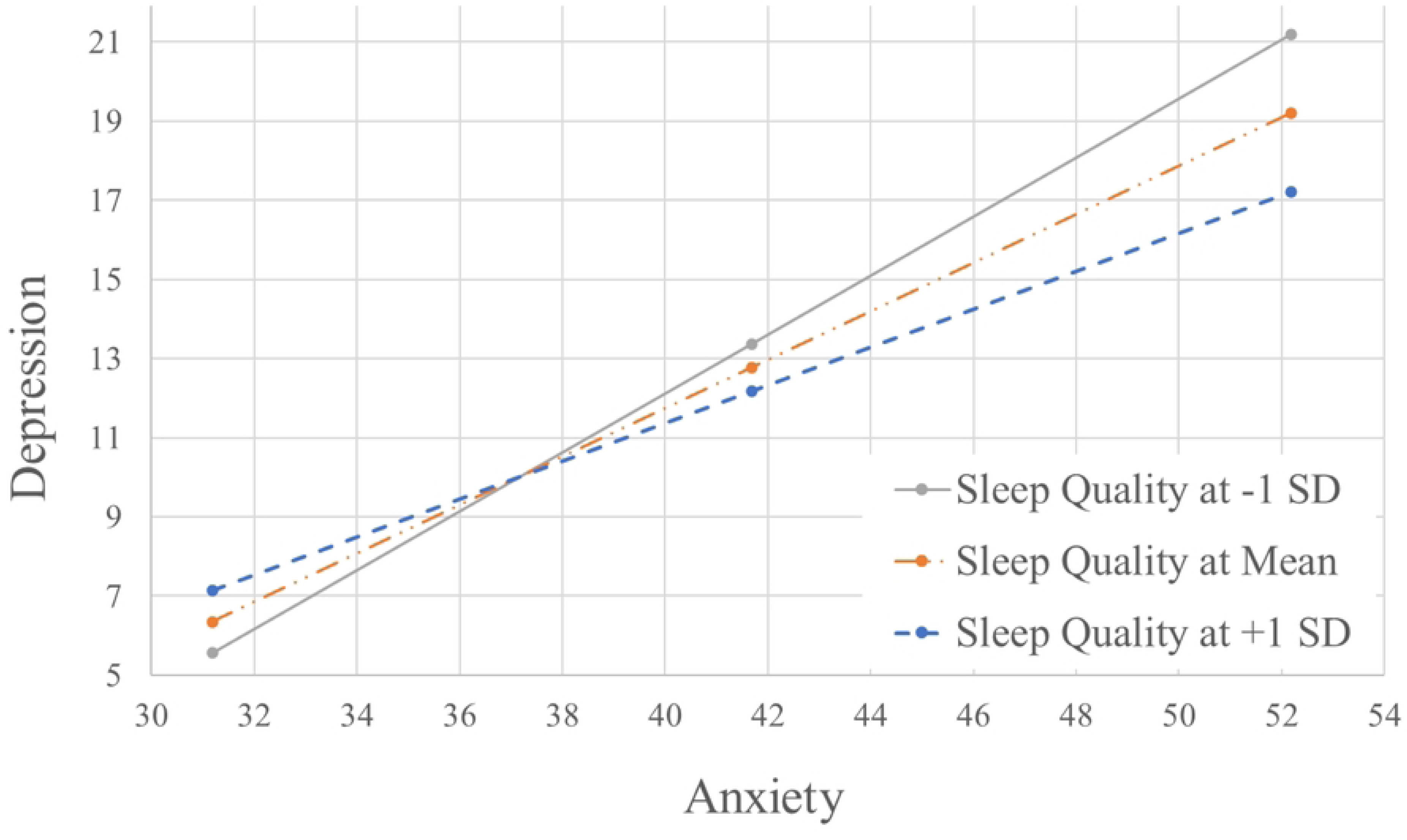
**Conditional Effects of Anxiety on Depression at Varying Levels of Sleep Quality**

## Discussion

This research explored the relationships among undergraduate stress, anxiety, and depressive symptoms, with a particular focus on anxiety as a mediating variable in the link between undergraduate stress and depressive symptoms, and on sleep quality as a moderator across all pathways in this mediation model. The following sections discuss the key findings and their implications for theoretical frameworks, intervention strategies, and directions for future research.

### Primary results

Supporting both previous works and Hypothesis 1, over 60% of first-year students disclosed having impaired sleep quality [77, 78], highlighting the need for a deeper understanding of its critical role in students’ adjustment to university life. Moreover, consistent with the literature and Hypothesis 2, students with reduced sleep quality reported increased anxiety [26], more depressive symptoms [22], and higher stress levels [28]. These findings support previous studies and extend current understanding by demonstrating the pervasive nature of poor sleep quality and its strong associations with mental health outcomes, particularly in the context of undergraduate stress.

Hypothesis 3 examined whether, as observed in general stress–depression relationships [12–17], anxiety could serve as an underlying mechanism linking undergraduate stress to depression. As anticipated, anxiety significantly mediated this relationship, indicating its role as a key mechanism through which undergraduate stress contributes to depressive symptoms. However, the direct effect persisted, suggesting that in a stressful undergraduate context, some students experience depressive symptoms in the absence of marked emotional distress such as anxiety, fear, or persistent worry. This supports the view that depressive symptoms may occur both independently of anxiety and in conjunction with it [79], challenging simpler models of direct stress–depression pathways [40, 80].

While initial results confirmed and extended previous insights into stress-related psychological distress, this study’s main objective was to investigate the interplay between undergraduate stress, anxiety, depression, and sleep quality using a moderated mediation model.

### Findings from the moderated mediation analysis

The moderated mediation analysis showed that, in the context of undergraduate stress, poor sleep quality significantly increases susceptibility to depression among students who also report elevated anxiety. By contrast, sleep quality did not moderate the direct path from stress to depression, nor the first segment of the indirect path from undergraduate stress-to-anxiety.

#### Comparison with mediation based findings

Sleep quality plays a central role in psychological well-being and has been identified as a mediating factor between stressful experiences and emotional distress. Specifically, recent research has shown that it mediates the associations between stress and anxiety, stress and depression, and anxiety and depression [35–37]. In these studies, sleep quality is treated as a downstream factor, which may encourage intervention approaches that primarily focus on addressing sleep problems once they have emerged.

By contrast, this study positions sleep quality not as an outcome, but as a vulnerability factor associated with stress-related emotional difficulties—highlighting the importance of recognising poor sleep quality as an early marker of psychological risk, rather than a symptom to be addressed only once distress becomes more pronounced.

#### Extending and contextualising prior moderation models

The current findings align with and extend previous research that has conceptualised sleep quality as an amplifying factor in stress-related psychological distress [40, 41, 43]. Notably, however, few studies have statistically modelled this relationship using a moderated mediation approach with anxiety as a mediator and sleep quality as a moderator of the stress– depression association. To our knowledge, this framework has only been applied in the present research and Ho’s [43] investigation. While Ho’s study relied on a highly heterogeneous sample of Facebook users aged 15–49 years affiliated with an unspecified administrative organisation, this study builds on these findings within a more clearly defined and developmentally focused population: university students in emerging adulthood. This group faces both age-specific transitional challenges [3, 81] and stressors unique to the undergraduate academic context [82]. By narrowing the population focus, this study enhances the generalisability of Ho’s findings and underscores the role of sleep quality as a potential amplifier of emotional distress during a particularly vulnerable developmental window [43].

The present study also supports the results reported by Park et al. [41], who found no evidence that sleep disturbance moderates the relationship between stress and anxiety. Replicating this outcome in a student sample strengthens the view that, while poor sleep quality contributes to emotional difficulties, it does not appear to affect whether anxiety emerges in response to stress. Rather, its influence seems to lie in intensifying depressive symptoms in individuals who are already experiencing anxiety [43, 44]. This is important, as the differentiation—between sleep disturbance’s lack of effect on anxiety and its role in amplifying anxiety-related depression—enhances the understanding of symptom-specific risk patterns in stress-related psychological distress.

While the present findings help clarify the distinct role of sleep disturbance in the indirect pathway between undergraduate stress and depression, they also diverge from earlier research suggesting that poor sleep quality exacerbates the relationship between general life stress and depressive symptoms [40, 41]. Several potential explanations may account for this discrepancy. First, this study used a moderated mediation model, while earlier studies [40, 41] employed a simple moderation approach that examined how sleep quality affects depressive symptoms related to stress without accounting for the mediating role of anxiety. This analytical difference may have led to an overestimation of the direct impact of sleep quality on the stress– depression relationship, potentially obscuring its more pronounced interaction with anxiety in predicting depressive symptoms. Second, sleep quality may interact differently with general life stress, as assessed in Leggett et al. [40] and Park et al. [41], than with undergraduate stress, which served as the focal stressor in this study. The type and source of stress could meaningfully shape how sleep quality functions as a moderator in these relationships. Third, age differences between samples may also help explain the divergent findings. The current sample, with a mean age of 21 years, differed substantially from the samples in Leggett et al. [40] and Park et al. [41], which had mean ages of 47 and 39 years, respectively. This age gap raises the possibility that individuals in emerging adulthood—such as first-year university students—may possess greater psychological adaptability or developmental resources to cope with poor sleep under stress than adults in their thirties or forties. Nonetheless, these interpretations remain speculative and indicate that more research is needed to clarify how developmental stage, stressor type, and sleep quality jointly contribute to the onset of depressive symptoms. Such research would enhance the understanding of age-specific vulnerability factors and inform the development of more finely tailored, developmentally sensitive prevention strategies.

### Implications for interventions

Within a moderated mediation framework, the findings suggest that reduced sleep quality amplifies the impact of undergraduate stress-related anxiety on depressive symptoms. This highlights the need for proactive, personalised support targeting early signs of anxiety and disturbed sleep, given that their co-occurrence is associated with elevated depressive symptoms. To translate these findings into practice, universities should implement evidence based strategies at both individual and structural levels. These may include sleep hygiene education and cognitive–behavioural therapy [83, 84], mindfulness-based interventions [85, 86], progressive muscle relaxation [87], peer support programmes [88], and technology-assisted approaches [89]. In addition, potentially stress-inducing academic conditions should be critically examined and revised where necessary—such as rigid curricula, excessive workloads, overcrowded classes, inefficient scheduling, and unclear academic expectations. Combining individual-level support with structural improvements can help create a learning environment that promotes psychological well-being, particularly among students at increased risk of depressive symptoms due to stress-related anxiety and poor sleep quality.

### Study limitations and directions for future work

Although this study advances the understanding of the risk factors for depression among stressed undergraduate students, some limitations warrant consideration in future research. First, owing to its cross-sectional nature, this study cannot establish cause-and-effect relationships. Although the results indicate that, in a stressful undergraduate context, poor sleep may heighten vulnerability to anxiety-related depressive symptomatology, alternative explanations are possible. For example, undergraduate stress might initially trigger emotional distress, with disrupted sleep emerging as a downstream effect. To confirm the validity of the directional relationships proposed in this study, future research should utilise longitudinal designs.

Second, although a retrospective power analysis showed that this study had adequate power (100%) to detect effects of the size observed, the modest sample of 102 participants may still have limited the ability to detect smaller or more subtle associations—especially regarding the lack of a significant moderating effect on the direct link between undergraduate stress and depressive symptoms. These effects may exist, but were not statistically detectable within the current sample. Therefore, future studies should aim to replicate the investigations via a larger and more representative sample to enhance the reliability and generalisability of the results.

Third, this study relies on self-report measures (USQ, PSQI, BDI, STAI-state), which, despite their widespread use, have known limitations, including social desirability bias, response tendencies, and limited objectivity. In particular, self-reported sleep quality and stress may not always align with physiological data, as some students might inaccurately perceive their sleep or stress levels (e.g., experiencing fragmented sleep despite feeling rested or elevated physiological stress even though feeling calm). Notably, though, objective measures alone may have limitations, as they may not provide insight into a person’s internal states and perceptions, which are also important indicators of well-being [90]. Future research should combine self-report and objective assessments, such as actigraphy or polysomnography for sleep quality, and biochemical or physiological measures for stress, to provide a more comprehensive evaluation.

Another constraint of this study is its use of total scores for sleep quality and undergraduate stress, which may obscure the effects of individual PSQI subcomponents and USQ items on anxiety and depression. However, future investigations could build on these findings by further exploring which subscores—or combinations thereof—are most strongly associated with anxiety-related depressive symptoms. For instance, different domains of undergraduate stress, such as academic pressure, financial strain, and personal or social stressors, may relate differently to mental health outcomes. Similarly, specific components of the PSQI—such as difficulties falling asleep, sleep disturbances, or daytime fatigue—might show stronger associations with psychological distress than the global sleep score alone. These insights can aid in the design of more tailored student support strategies that address the most impactful stressors and sleep difficulties.

Beyond methodological refinements, various demographic, social, psychological, and physiological factors may influence how students respond to stress and their vulnerability to associated mental health challenges such as anxiety and depressive symptoms. As the present findings are based primarily on a sample of young, female psychology students in their first year of study, they may not be generalisable to other student populations—particularly those differing in age, sex, field of study and study tenure. Other important factors not captured in this study may include cultural or socioeconomic background, mental health history, family circumstances, and immigration status. To better account for this heterogeneity, further investigations should examine these relationships in more diverse student populations to gain nuanced insights into individual risk profiles and develop targeted prevention strategies tailored to specific groups.

## Conclusion

This study highlights the high prevalence of poor sleep quality among first-year university students and its strong association with undergraduate-specific stress, anxiety, and depressive symptoms. Moreover, the data supported a statistical mediation model in which anxiety linked undergraduate stress to depressive symptomatology—an association that, to date, has rarely been examined using stressors specifically tied to the undergraduate context.

The central findings concern the role of sleep quality in influencing the psychological outcomes associated with undergraduate stress: Sleep difficulties amplified the relationship between anxiety and depressive symptomatology, such that the indirect effect of undergraduate stress on depression was most pronounced among students experiencing both high anxiety and poor sleep. In contrast, sleep quality did not moderate the association between undergraduate stress and anxiety, nor the direct link between undergraduate stress and depressive symptoms. These patterns point to a distinct function of sleep quality—not as a general risk factor, but as a targeted amplifier of anxiety-related depressive symptoms in this population.

Together, these findings expand current knowledge of psychological distress in the undergraduate university setting and illustrate how individual differences—particularly in sleep quality—shape the associations among undergraduate stress, anxiety, and depression. From a preventive perspective, the early identification of students with co-occurring elevated anxiety and poor sleep quality appears essential, as this combination is associated with an increased risk of depressive symptoms. Targeted interventions that address both anxiety and sleep difficulties may be particularly effective in mitigating mental health problems during the vulnerable transition into university life.

## Acknowledgements

This manuscript is based on doctoral research conducted at the University of Westminster. As the lead author is not a native English speaker, AI tools including ChatGPT, Claude, and Grammarly were used to support language editing and improve clarity and flow. The final manuscript was also professionally edited by Editage. The content, interpretations, and analyses are entirely the authors’ own. The authors also acknowledge Editage (www.editage.com) for assistance with manuscript preparation and submission.

## Data availability statement

The data underlying this study consist of anonymised survey responses measuring undergraduate-specific stress, anxiety, depressive symptoms, and sleep quality. All relevant data will be made publicly available in OSF upon the acceptance of the manuscript. During peer review, the data can be shared with the editor or reviewers upon request.

## Supporting information

**S1 File**. Assumption testing workbook. Excel file containing residual plots, normality tests, multicollinearity diagnostics, and other assumption checks across multiple sheets.

**S2 File**. STROBE checklist. Completed checklist outlining adherence to the Strengthening the Reporting of Observational Studies in Epidemiology (STROBE) guidelines.

## Notes

### Competing Interest Statement

The authors have declared no competing interest.

### Funding Statement

The author(s) received no specific funding for this work.

### Author Declarations

Approval for this research was granted by the Ethics Committee of the University of Westminster (reference number: ETH1617-1048).

